# Sepsis in patients with vs. without mental illness: a comparison of demographic, insurance, comorbidity, and infection source characteristics

**DOI:** 10.1101/2024.04.15.24305016

**Authors:** Yi-Ru Chen, Melva Morales Sierra, Jaime Jacob, Lisa Iyeke, Lindsay Jordan, Khatija Paperwalla, Mark Richman

## Abstract

**Background:** Adherence to the Surviving Sepsis Campaign’s 3- and 6-hour bundles (blood cultures/serum lactate/antibiotics/IV fluids/vasopressors) improves mortality. Septic patients with mental health illness may not receive optimal care, being unable to explain symptoms, understand/accept their condition/care, or remain calm. We compare characteristics of ED septic patients with vs without mental health illnesses in their demographics, insurance, housing status, comorbidities, and infected organs, part of a larger, retrospective study seeking to compare such patients’ sepsis care quality (bundle adherence, length-of-stay (LOS)).

**Methods:** Patients with sepsis (and their infection source) between 2017-2019 were identified from a health system sepsis reporting database. Their characteristics of history of mental health illness (cognitive, mood, personality, schizophrenia, psychosis, substance use), demographics, insurance, housing status, and comorbidities were obtained via electronic health record query. Such characteristics were compared among patients with vs without mental illness.

**Results:** A greater percent of septic patients than the overall U.S. population had a mental illness (26.9% vs 21%). In univariate analysis, patients with mental illnesses were older (77.9 vs 67.6 years), more-likely to be from a psychiatric facility/group home (32.5% vs 2.1%; p<0.0001), have Medicare (58% vs 47.1%; p=0.0201), have ≥1 comorbidity (72.6% vs 0.6%; p<0.0001), and have a respiratory source of sepsis. People without mental illnesses were more-often from either a private home/nursing home (75.2% vs 56.7%; p<0.0001) or clinic (8.9% vs 3.2%; p=0.0191), have private insurance (15.2% vs 7.6%; p=0.0167), and have immune-modifying medications or cancer (20.7% vs 7%; p<0.0001).

**Discussion:** Compared with national rates of mental health illness, a higher percent of septic patients (26.9% vs. 21%) had mental illness (mostly neurocognitive). Septic patients with mental illnesses tended to be older, from a psychiatric facility/group home, have Medicare, have ≥1 comorbidity, and have a respiratory source of sepsis. Patients without mental illnesses more-often had immune-modifying medications and malignancy. Patients with mental health illnesses were more-likely to have a respiratory source of their sepsis, perhaps due to higher risk for aspiration or respiratory contagion in group homes/psychiatric facilities. Clinicians may want to specifically test or empirically treat for respiratory sources in this population. Future analyses (multivariate) will determine whether differences in quality of care, mortality, or length of stay exist, and if specific characteristics above were associated with these different outcomes.

**Limitations:** The study was based on data from a single hospital system, which might limit generalizability. Additionally, the study relied on data collected for sepsis management, which might not capture all patients with sepsis or those who did not receive the sepsis bundle. Furthermore, all data collected pertaining to this study was only in the timeframe between 2018-2019. The proportion of septic patients with vs. without mental health disorders during and following the COVID pandemic may differ from the earlier time frame represented in this study.

## Introduction

Sepsis is a major health problem, with increasing prevalence, high costs, and poor outcomes. In the United States, sepsis accounts for 1 million hospitalizations and 5.2% of hospital expenditures (>$20 billion/year).^12^ Sepsis accounts for 25–50% of in-hospital mortality; 6-month mortality of post-discharge patients ranges from 45% to 65%.^1^ If patients with these conditions survive, they have increasing chronic complications, morbidity, high costs of care, and decreasing quality of life.^6,13^ Many of the over 1.5 million annual cases of sepsis in the United States present to the Emergency Department (ED).^4^ Prompt identification of sepsis and treatment with broad-spectrum antibiotic agents and intravenous fluids,^2, 5^ as recommended by clinical practice guidelines and the Centers for Medicare and Medicaid Services (CMS’s) Surviving Sepsis Campaign,^8,9^ produces improved outcomes, including decreased mortality.

Sepsis is defined as having ≥2 SIRS criteria (temperature >38.0°C or hypothermia <36.0°C, heart rate >90 beats/minute, respiratory rate >20 breaths/minute, leukocytosis >12*10^9^/L or leukopenia <4*10^9^/L) and a likely bacterial source of their SIRS. Patients meeting these criteria should, within 3 hours of sepsis being suspected, have:

1. Serum lactate measured.
2. An intravenous fluid bolus (IVF) of at least 30 mL/kg for patients with elevated lactate or hypotension (mean arterial pressure (MAP <65 mmHg), unless contraindicated by evidence or risk of volume overload (eg, congestive heart failure, dialysis)
3. Vasopressors initiated in fluid-non-responsive patients with elevated lactate or hypotension
4. Blood cultures drawn before antibiotics are given
5. Broad-spectrum antibiotics administered

Additionally, among those with initially-elevated lactate, a repeat serum lactate (should be performed within 6 hours of sepsis being suspected, to determine whether tissue perfusion improved with IVF and vasopressors

Patients may not receive the complete bundle for a variety of reasons (eg, delay in recognizing sepsis, contraindication to the full 30 mK/kg IVF bolus, or an advanced directive limiting use of bundle interventions).

Previous research has demonstrated patients with severe mental illness and cardiovascular disease or diabetes are 30% less likely to receive recommended cardiovascular or diabetes care, including hospitalization, diagnostic tests, appropriate medications, and invasive procedures.^10^ It is possible such disparities apply to severe sepsis or septic shock, as well. However, very few such studies have been performed. A national cross-sectional study in Sweden found patients with severe mental illness had increased risk of death associated with influenza/pneumonia (OR = 2.06) and sepsis (OR = 1.61). They also had an increased risk of hospitalization associated with influenza/pneumonia (OR = 2.12) and sepsis (OR = 1.89), suggesting greater severity.^7^ This current study aims to add to the literature regarding mental illness and sepsis outcomes.

Patients with a severe mental illness crisis may not get full-bundle care for several reasons. They might: 1) be currently psychotic or manic and unable to explain their symptoms appropriately, 2) not understand their condition or the care they’re offered, 3) refuse care, 4) be agitated or aggressive, or 5) take medications whose side effects could be misinterpreted as sepsis (eg, neuroleptic malignant syndrome (NMS)). We hypothesize that patients with a concurrent severe mental health crisis will have increased risk of not receiving the full CMS sepsis bundle compared with those without concurrent severe mental health crisis.

The current study compares characteristics of septic patients with vs. without an acute mental illness in the realms of demographic, insurance, comorbidities, and infection source characteristics. This is part of the above-mentioned broader and larger study our group is conducting aiming to determine whether having a severe mental health crisis impacts the quality and timeliness of sepsis bundle care.

## Methods

Northwell Health is a 22-hospital health system largely operating in Long Island and New York City. Long Island Jewish Medical Center (LIJMC) is a 583-bed tertiary care teaching hospital serving a racially and socio-economically diverse population. The hospital is 3 miles from the 349-bed, 20-story Creedmoor National Psychiatric Center and its multiple associated group homes and clinics, and is a receiving center for Creedmoor patients. The adult ED sees approximately 100,000 patients per year. Members of LIJ’s Department of Quality Services collect and submit sepsis data to state and federal agencies. Patients with a contraindication to bundle components (pregnant or those with advanced directives for non-aggressive medical care) are removed from the sepsis database.

All ED patients meeting sepsis criteria on the basis of vital signs and suspected infection qualify for the ED sepsis database. LIJMC ED has a “sepsis trigger” system whereby if patients meet vital sign or WBC lab criteria for sepsis, the patient’s row on the ED electronic board is changed to red. During the daytime, there is active surveillance by “sepsis champions.” If they identify a patient meeting sepsis criteria who are not being managed according to “Surviving Sepsis” guidelines, the patient’s providers are notified and encouraged to use the “Sepsis Order Set.” The ED has, within it, a separate, locked area for patients with behavioral health (BH) disorders; all patients get a medical evaluation by a Board-certified Emergency Physician, in addition to an evaluation by a Psychiatrist. Patients with psychiatric illness who are suspected of having sepsis (either at triage or during psychiatric evaluation) on the basis of vital signs, infectious symptoms, or concerning lab values) are either triaged to the “main/medical” area of the ED (not to the BH area), or are moved from the BH area to the “main/medical” area of the ED. With these multiple levels of screening and intervention. Missing cases of sepsis are rare; nearly every patient meeting sepsis criteria, including those with severe psychiatric illness, have a complete blood count, lactic acid, urinalysis, urine culture, blood cultures, and a respiratory viral panel.

Patients were included in this study if they:

1. Attended the LIJ adult ED between December 2017 and December 2019, and
2. Were in the LIJ Department of Quality Service’s sepsis database, and
3. Had any of the ICD-10 codes in Appendix A for mental health conditions in the broad categories of: 1) dementia (including Alzheimer’s disease), 2) other neurocognitive disorders, 3) mood disorders (eg, depression), 4) personality disorders (eg, borderline) 5) psychotic disorders (eg, schizophrenia), or 6) substance use disorders (eg, alcohol).

We reviewed the chart for the following domains:

1. Demographic data: age, gender, race/ethnicity, and insurance (Medicare, Medicaid, commercial, none/self-pay)
2. Where patient arrived from: individual private home, group home, community psychiatric residence/apartment, psychiatric facility, transfer from other hospital, transferred from skilled nursing facility or nursing home, police custody
3. Past medical history/comorbidities (eg, congestive heart failure, chronic obstructive pulmonary disease, immune suppression)
4. Infection source

This project was deemed to fall under the realm of performance improvement by the Institutional Review Board.

## Results

Five hundred eighty three patients with sepsis were identified; of these 157 had one of the above-mentioned mental health disorders and 426 did not. (**Figure 1**) A greater percent of septic patients had a mental illness (26.9%) than the overall U.S. population (21.0%)^11^ and than LIJ ED’s non-septic population (5.8%) (p < 0.0001). There were no differences in the percent of patients with vs. without mental illness in concerning vital signs: temperature ≥ 38.0 C (p = 1.0), heart rate ≥ 110 beats per minute (p = 0.51), systolic blood pressure <100 mmHg (p = 0.25), room air oxygen saturation <95% (p = 0.91), respiratory rate ≥20 per minute (p = 0.85) (**Table 1**); nearly 78% of septic patients with mental illness did not have a fever.

**Figure 1.**
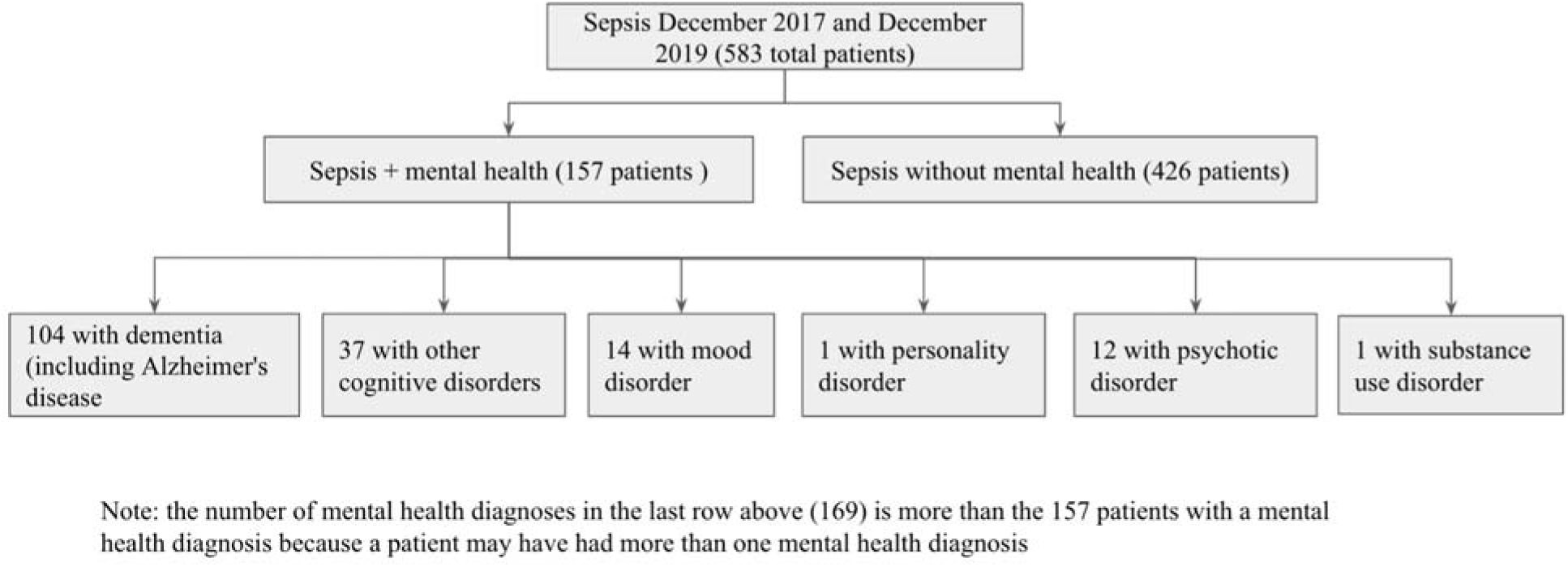
CONSORT Diagram

**Table 1.** Concerning Vital Signs.

| Abnormal Presenting Vital Signs | History of mental illness (%) | No history of mental illness (%) | P value |
| --- | --- | --- | --- |
| temperature $\geq 38.0$ C | 22.3 | 22.8 | 1.0 |
| heart rate $\geq 110$ beats per minute | 34.4 | 37.3 | 0.51 |
| systolic blood pressure $<100$ mmHg | 36.3 | 31.2 | 0.25 |
| room air oxygen saturation $<95\%$ | 24.8 | 24.4 | 0.91 |
| respiratory rate $\geq 20$ per minute | 40.7 | 39.9 | 0.85 |

In univariate analysis, patients with mental illnesses were older (77.9 vs 67.6 years), had Medicare (58.0% vs 47.1%; p = 0.02) (**Figure 2**), were more-likely to be from a psychiatric facility/group home (32.5% vs 2.1%; p < 0.0001) (**Figure 3**), have at least one comorbidity (72.6% vs 0.6%; p < 0.0001) (**Table 2**), and have a respiratory source of sepsis (47.1% vs. 38.3%; p < 0.04) (**Figure 4**). Among septic patients with mental health disorders, approximately 83% had dementia (often concurrent with delirium), 10% had psychosis, and 6% had bipolar disorder. Patients without mental illnesses more-often arrived from either a private home/nursing home (75.2% vs 56.7%; p < 0.0001) or clinic (8.9% vs 3.2%; p = 0.0191), have private insurance (15.2% vs 7.6%; p = 0.0167), have immune-modifying medications or cancer (20.7% vs 7.0%; p < 0.0001), and have a urinary tract source of sepsis (36.9% vs. 24.0%; p < 0.02).

**Figure 2.**
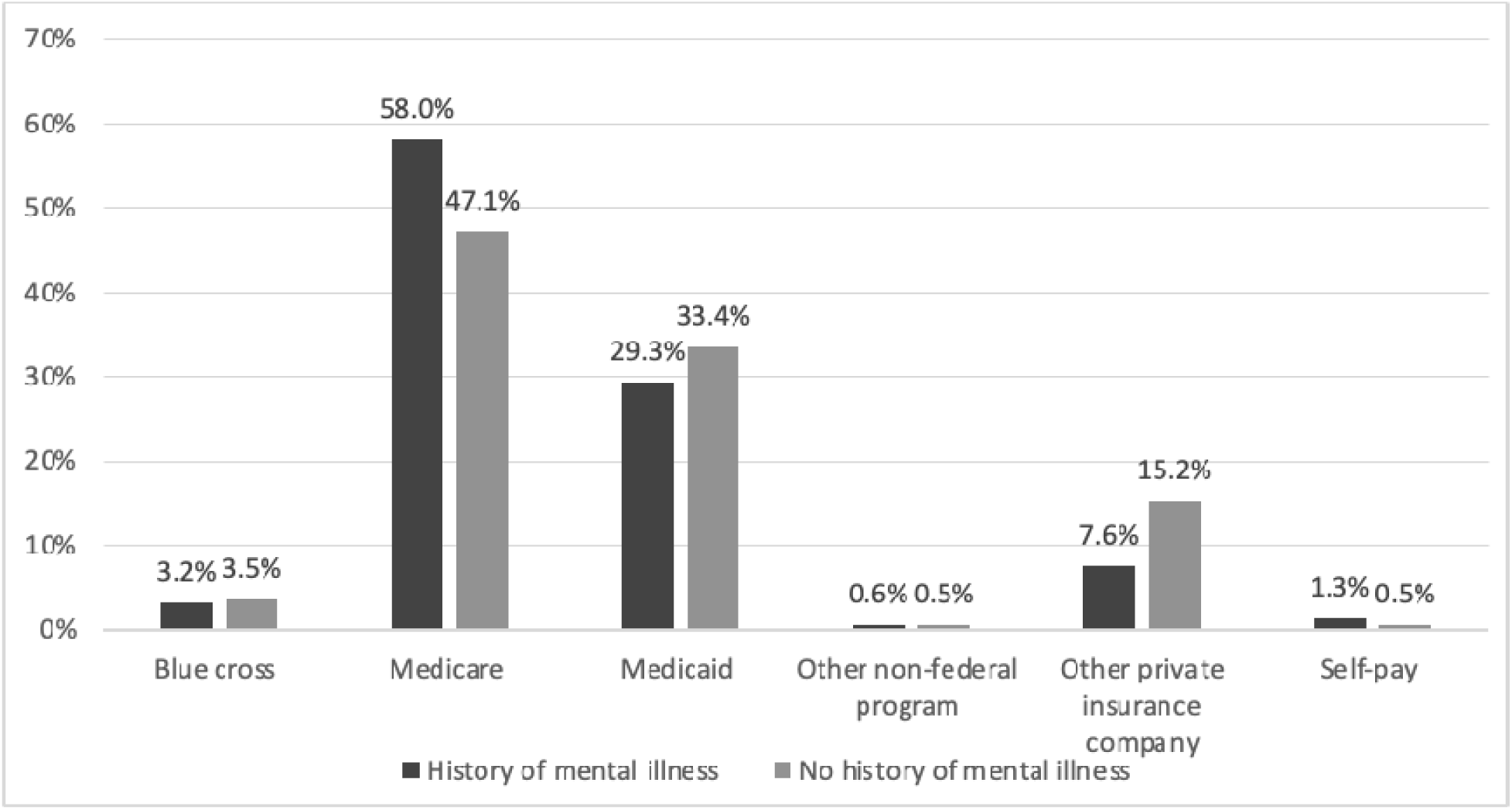
Source of Health Insurance

**Figure 3.**
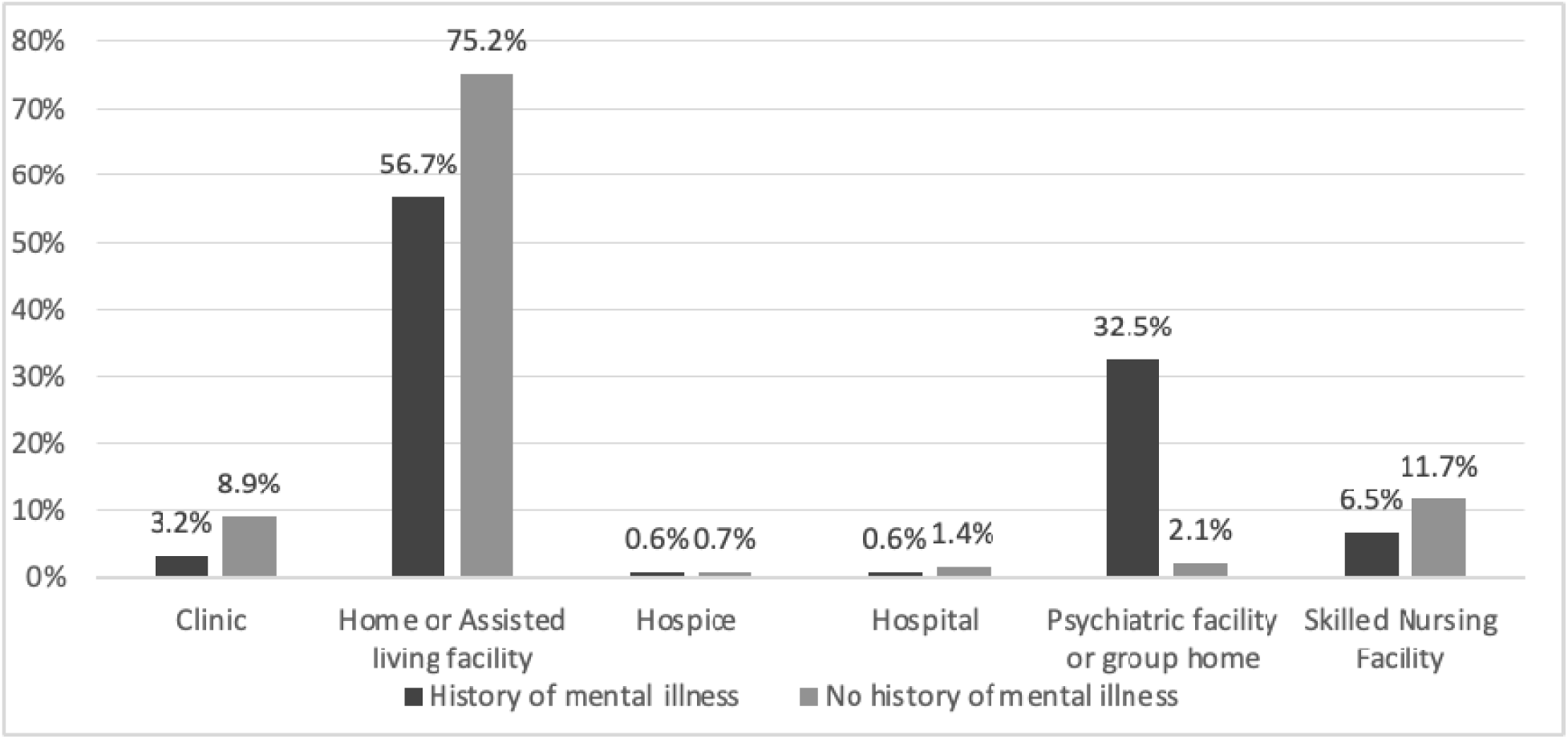
Source of Admission

**Figure 4.**
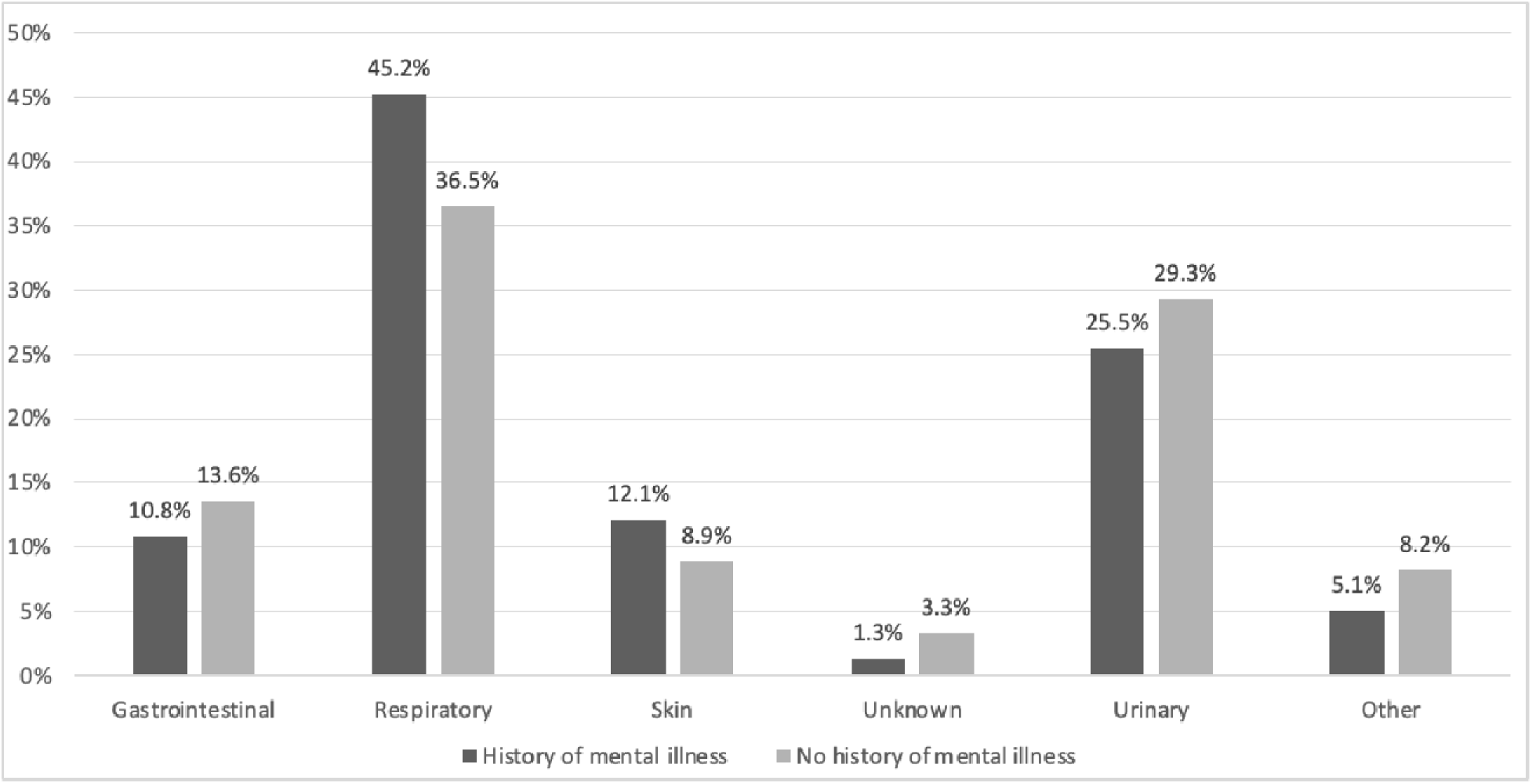
Source of Sepsis in Organ Systems

**Table 2.** Comorbidities.

| Comorbidity | History of mental illness (%) | No history of mental illness (%) | P value |
| --- | --- | --- | --- |
| AIDS or HIV | **3.8** | 0.5 | **<0.01** |
| Altered mental status | **70.1** | 43.2 | **<0.0001** |
| Chronic liver disease | 0.6 | 2.6 | 0.13 |
| Chronic renal failure | 6.4 | 8 | 0.52 |
| Chronic respiratory failure | 0 | 0.5 | 0.39 |
| Congestive heart failure | 19.1 | 15 | 0.24 |
| Diabetes | 40.6 | 43.3 | 0.58 |
| Immune-modifying medications | 7 | **20.7** | **<0.01** |
| Lower respiratory infection | 45.2 | 46.5 | 0.78 |
| Lymphoma, leukemia, or multiple myeloma | 0.6 | **5.9** | **<0.01** |
| Metastatic cancer | 3.8 | **12.7** | **<0.01** |
| Organ transplant | 0.6 | 2.1 | 0.21 |
| ≥1 comorbidity | **72.6** | 60.3 | **<0.01** |

## Discussion

Mental illness was more prevalent among LIJ ED patients with sepsis (26.9%) than among the broader U.S. population (21.0%^11^) and than among our ED’s non-septic population (5.8%; p <0.0001). Such results are consistent with the above-mentioned Swedish study’s^7^ observation that patients with mental health disorders were more likely to have sepsis.

Septic patients with mental illnesses tended to be older, from a psychiatric facility/group home, have Medicare, and have a respiratory source of sepsis (while patients without mental illness more-often had a urinary tract source of sepsis). Most sepsis patients had more than one comorbidity; those with mental illness were more-likely than those without, and were more-likely to have HIV/AIDS (though this rate was low), whereas those without mental illness were more-likely to have malignancy or immune-modifying medications. There was no difference in rates of diabetes.

The higher rate of respiratory source of sepsis among those with mental illnesses may be due to a higher risk for aspiration owing to dementia, or contagion of respiratory pathogens in group homes or psychiatric facilities.^4^ Clinicians may want to specifically test or empirically treat for respiratory sources in this population. Such patients also more-often had skin infections, possibly owing to poor personal or environmental hygiene.

There was no difference between septic patients with vs. without mental illness in likelihood of having any particular abnormal vital sign. Nearly 78% of septic patients with mental illness did not have a fever. ED behavioral health and non-behavioral health clinicians should be vigilant for infectious conditions in this population (which may exacerbate their behavioral health problems), without relying exclusively on fever as a marker of infection.

Research in Sweden using a large national registry compared the demographic characteristics and sepsis outcomes of people with vs. without serious mental illnesses (psychotic, bipolar, and single manic episodes).^8^ Patients with mental illness were more-likely to be middle-aged (40-69 years),while those without mental illness were more often older. The authors caution that fewer older patients might have had mental illness because those with severe mental illness might have perished prior to reaching old age. People without mental illness were slightly less-likely to have cardiovascular and pulmonary comorbidities. In comparison, our study found patients with mental illness were more-likely to be older and have non-infectious immune disorders and cancer.

## Limitations

The study was based on data from a single hospital system, which might limit generalizability. Furthermore, all data collected pertaining to this study was only in the timeframe between December 2017-December 2019. The proportion of septic patients with vs. without mental illness during and following the COVID pandemic may differ from the earlier timeframe represented in this study.

## Conclusion

Significant demographic differences exist between septic patients with vs. without mental illness. In addition, clinically-significant differences exist in that the source of infection was more-often respiratory among patients with mental illness and urinary among those without. Future analyses (multivariate) will determine whether differences in quality of care, mortality, or length of stay exist between patients with vs. without mental illness, and if specific patient characteristics were associated with these different outcomes.

## Data Availability

All data produced in the present study are available upon reasonable request to the authors.
All data produced in the present work are contained in the manuscript.

## Acknowledgements

None

**HLPRS - Hofstra Longitudinal Pre-medical Introduction to Research Society:**

Zebin Zinia, Aman Umer, Zoya Ramzan, Ujala Nigam, Sadia Mebin, Rochelle Hall, Casey Urban, James Petrancosta, Priyanka Mehta, Jeffrey Hoffman, Valeria Gromova, Stephanie Fink, Saher Chaudhry, Sarah Boladale Johnson, Jeffrey Wang, Prashanth Thomas, Komal Khokhar, Jun Ha Joung, Gissele Cardenas, Demetra Menoudakos, Adeola Taiwo, Laiba Sandhu, Genelle Ramjattan, Gisette Ochoa, Chinna Njoku,Ruchika Khindri, Vicky Zhu, Zarina Davies, Annie Wang, Saher Shafiq, Trey Rogers, Jaime Jacob, Stephen Guilherme, Niralee Rana, Victoria Gaylor, Christian Dietz, Adam Desouky, Esther Gestetner, Cristina Martinez, Matthew Chu, Sarah Real, Prince Multani, Alma Glavatovic, Megan Carroll, Naya Mahabir, Julia Razzante, Melanie Heney, Samuel Brown-White, Sean Kane, Retu Domnic, Aliya Catanzarita, Tarek Harhash, Abigail Jara, Aakriti Sharma, Yi-Ru Chen, Shaina Silverman, Rida Nasir, Grace Sanker, Daniel Ehling, Melva Morales Sierra, Sophia Trojanowski, Alexandria Slusser, Angelina Ryba, Louis Robustelli, Mohammad Rahman, Sandhya LoGalbo, Jasmine Kumar, Jasleen Khatra, Romesha Khan, Mark Khan, April Ivan, Brendan Colas, Simranjit Athwal, Ravesh Bechu, Brenda John, Martina Lim, Yashwinder Kaur, Akshita Sawani, Tejbir Singh, Zuhair Ishraq

## Appendix A

### ICD-10 codes for severe mental illness

| Neurocognitive Disorders |
| --- |
| Alzheimer's (G30.0) |
| Alzheimer's late onset (G30.1) |
| Alzheimer's specified NEC (G30.8) |
| Alzheimer's with behavioral disturbance (G30.9) |
| Delirium (F05) |
| Dementia (F03.90) |
| Dementia with aggressive behavior (F03.91) |
| Frontal Dementia (G31.09) |
| Hepatolenticular degeneration (E83.01) |
| HIV infection (F02 and B20) |
| Huntington's disease (F02 and G10) |
| Lewy bodies (G31.83) |
| Multiple etiologies (F03) |
| Neurosyphilis (A52.17 ) |
| Parkinson's with behavioral disturbance (F02 and G20) |
| Senile with acute confusional state (F05) |
| Wilson's disease (E83. 01) |
| Vascular (acute onset) (mixed) (multi-infarct) (subcortical) (F01.50) |
| Vascular (acute onset) (mixed) (multi-infarct) (subcortical) with behavioral disturbance (F01.51) |
| Vitamin B12 deficiency (D51.9) |
| **Mood Disorders** |
| Bipolar disorder (F31.*) |
| Major depression (F32.0) |
| **Personality Disorders** |
| Antisocial personality disorder (F60.2) |
| Borderline personality disorder (F60.3) |
| **Psychotic Disorders** |
| Catatonia (F06.1) |
| Delusional disorder (F22) |
| Schizoaffective disorder, bipolar type (F25.*) |
| Schizophrenia, paranoid (F20.0) |
| Schizophrenia, disorganized (F20.1) |
| **Substance Use Disorders** |
| Alcoholic (F10.97) |
| Alcoholic with dependence (F10.27) |
| Alcohol withdrawal (F10.239) |
| Cocaine use, unsp with unspecified cocaine-induced disorder (F10240) |
| Inhalants with dependence (F18.27) |
| Psychoactive drug (F19.97) |
| Sedative/hypnotic/anxiolytic dependence w withdrawal, unsp (F13.239) |
| Sedatives, hypnotics or anxiolytics (F13.97) |
| Sedatives, hypnotics or anxiolytics with dependence (F13.27) |
| Volatile solvents (F18.97) |

